# Addressing Acute Febrile Illness Using a Syndromic Approach During A Chikungunya Epidemic in Rio de Janeiro, Brazil: A Prospective Observational Study

**DOI:** 10.1101/2023.04.15.23288370

**Authors:** José Moreira, B. Leticia Fernandez-Carballo, Camille Escadafal, Sabine Dittrich, Patrícia Brasil, André M Siqueira

## Abstract

**Background:** Identifying etiologies of acute febrile illness (AFI) is challenging in settings with limited laboratory capacity. We aimed to describe the causes of AFI among non-severe patients seeking care at the primary level in Rio de Janeiro, Brazil when a large chikungunya virus (CHIKV) epidemic was ongoing.

**Methodology/Principal Findings:** We conducted a 10-month prospective AFI study in participants aged 2-65 seeking care at public emergency departments and outpatient clinics. Patients with fever ≤ 7 days were offered enrollment, and clinical, and laboratory data were gathered for consecutive participants. A syndrome-driven approach comprising culture, molecular and serologic tests were adopted to investigate the cause of fever. Logistic regression model determined predictors of laboratory-positive CHIKV. Follow-up visits were conducted 14-28 days after the index visit. Five hundred participants (median age 26 [15-41] years, 50.4% females) yielded 824 diagnoses, and 249/500 (49.8%) of whom had multiple diagnoses. Systemic infection (382/500, 76%), followed by acute respiratory infection (155/500, 31%), and urinary infection (23/500, 4.6%) were the most common febrile syndromes. CHIKV was the primary etiology found in 284 (56.8%) participants. Viral upper respiratory infection accounted for 40/155 (25.8%) of the respiratory infections, of which Rhinovirus and Influenza A were the main viruses commonly detected. None of the diagnostic tests were positive in 124/500 (25%). Predictors of laboratory-positive CHIKV were the absence of cough, arthralgia, rash, high temperature, and leucopenia. Of those 297/500 (59.4%) who returned for the follow-up, 120/297 (40%) persisted with symptoms. CHIKV-positive patients were more likely to experience persistent arthritis than CHIKV negative [OR: 10.18 (3.64-28.45)].

**Conclusions/Significance:** Using a syndromic approach to identify the etiology of fever during an epidemic of CHIKV in Rio, we found evidence of other pathogens associated with AFI. Clinical and laboratory markers might allow early identification and accurate distinction of patients with CHIKV from other AFI to guide proper clinical management. Future research should assess whether a syndromic approach to febrile illness in resource-limited settings improves patient outcomes and rationale antimicrobial use.

Clinicaltrials.gov registration number: NCT03047642

## Introduction

The fever landscape across Latin America (LA) has changed profoundly in the last three decades, considering the significant reduction in malaria transmission in the region [1]. This epidemiologic shift has created a gap in knowledge about the causes of fever other than malaria, commonly referred as a non-malaria febrile illness - NMFI. As a clinical entity, NMFI represents a diagnostic challenge for the front-line clinicians, mainly when outbreaks of other acute febrile illness (AFI) occur, highlighting the vital role that diagnostics play in ascertaining the causative pathogen and appropriate supportive care [2]. Clinical management is often driven by syndrome-based guidelines employing empiric treatment [3,4]. In the absence of systematically collected data on fever etiology, a considerable mismatch between clinical diagnosis, clinical management, and actual etiology may occur, resulting in poor patient outcomes [5].

A recent review on the epidemiology of NMFI in LA identified only 17 studies in 8 countries conducted between 2007 and 2016 and found that the most frequently reported pathogens were dengue virus (DENV), followed by chikungunya virus (CHIKV), zika virus (ZIKV), Leptospira spp., Rickettsia spp., and Hantavirus spp. [6]. However, in this review, most studies lacked standardization and uniformity in study design and laboratory definitions and did not look for multiple fever causes.

Rio de Janeiro has been hardest hit by emerging and re-emerging arbovirus epidemics since the introduction of DENV in 1986 [7], CHIKV in 2014 [8], and ZIKV in 2015 [9]. The city has been struggling with a CHIKV epidemic and has registered an increase of ∼ 300% of the cases, compared to the same period in 2019 [10].

Here, a harmonized syndromic approach is incorporated into a more extensive study around fever host-biomarkers [11], to assess the etiology of AFI among non-severe patients seeking care at the primary level in urban Rio de Janeiro, Brazil, a time when a large CHIKV epidemic took place. We sought to describe comprehensively the primary febrile syndromes, the underlying etiologies, and the laboratory characteristics of febrile participants using conventional standard diagnostic techniques. We then highlighted the role of CHIKV as a cause of fever during the study period, describe its predictors and outcomes compared to CHIKV negative patients.

## Methods

This investigation was part of the Biomarker for Fever-Diagnostic (BFF-Dx) study [11]. Here, we describe the cause of fever in non-severe AFI patients seeking care at primary clinics and emergency departments in Rio de Janeiro, Brazil, between October 28, 2018, to July 31, 2019. Study registration on clinicaltrials.org (NCT03047642) was done before data acquisition, and further details about the study protocol have been described elsewhere [11]. We reported as per the Strengthening the Reporting of Observational Studies in Epidemiology (STROBE) guideline (see Supplementary appendix).

### Study site and population

Rio de Janeiro is located in Brazil’s southeastern macro-region, and the Atlantic Ocean limits it to the south (Figure 1). The estimated population for 2018 was 6.688.927 inhabitants (https://www.ibge/gov.br/). The city’s climate is tropical, hot, and humid, with local variations due to differences in altitude, vegetation, and proximity to the ocean. The average annual temperature between 1981 and 2010 was 29°C, with the highest daily temperature averages (from 30-32°C) occurring in the summer (March-December). The latter is a period in which the greatest precipitations are reported (i.e., an average of 205 mm). Despite being the second-largest city in Brazil, 20% of its inhabitants live in slums (also known as *favelas*), where access to necessary sanitary and water infrastructure is lacking, and high rates of violence occur [12].

**Figure 1.**
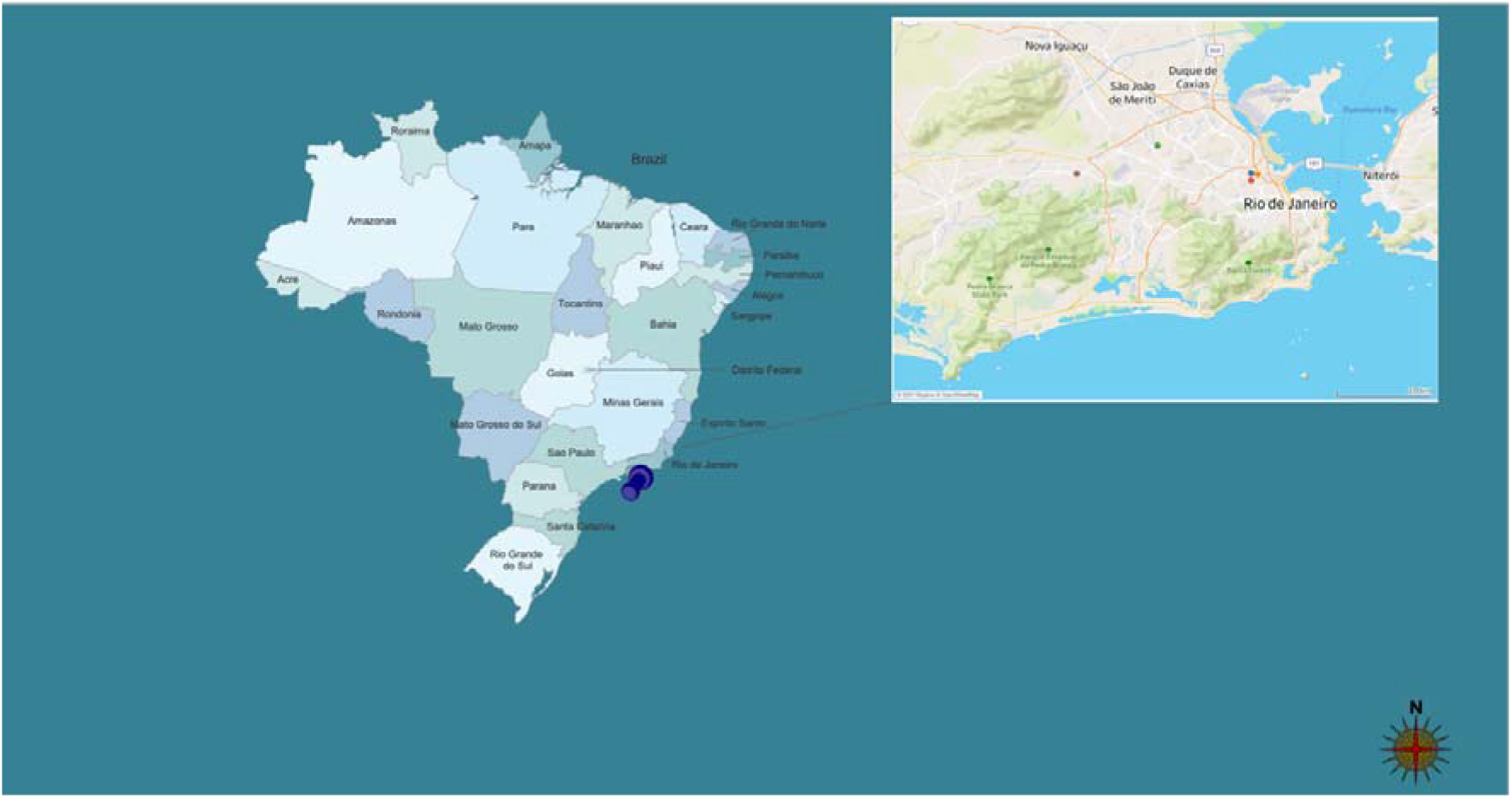
Location of study sites, acute febrile illness, Rio de Janeiro, October 2018-July 2019. Map showing the location of Rio de Janeiro state in Brazil. The inlet shows the location of study sites and coordinating center in urban Rio de Janeiro.

We recruited non-severe adults and children in two public emergency departments and outpatient clinics from two primary health care clinics, localized at the north and west part of the city during the study period. The mean number of monthly medical visits reported by each emergency department was 6.000 and served a population of about 80.798 individuals. The study coordinating center in Brazil was the Instituto Nacional de Infectologia Evandro Chagas (INI/Fiocruz) - a regional reference center for diagnosing and treating infectious diseases in Rio de Janeiro. This work received ethical approval from the National Review Board and (CONEP) and local review board (INI/Fiocruz), under the protocol number 70984617900005262.

### Clinical assessment

All patients were clinically managed according to local standards of care. Data was captured using paper and electronic standardized case-report form (OpenClinica Enterprise 34). A follow-up visit was scheduled between 14 to 28 days to perform fever outcome evaluation and laboratory assessments.

### Laboratory investigations

All laboratory investigations were done following standard procedures and have been previously described in detail elsewhere [11].

We performed a rapid diagnostic test (RDT) against ZIKV, DENV, and CHIKV using the DPP^®^ ZDC IgM/IgG (Bio-Manguinhos, Fundacao Oswaldo Cruz, Brazil) assay in acute-phase samples. In a random sample of the participants (*n* = 294/500, 58.8%), we performed quantitative reverse transcriptase-polymerase chain reaction for ZIKV, DENV, and CHIKV on acute-phase samples of patients with evidence of acute undifferentiated febrile illness [13]. Acute-phase samples were also tested by enzyme-linked immunosorbent assay (ELISA) for the presence of IgM and IgG antibodies against ZIKV (Euroimmune, Germany), DENV (Euroimmune, Germany), and CHIKV (Euroimmune, Germany) [14].

We adopted a symptom-driven approach to investigate the causes of fever in study participants [11]. All laboratory investigations were conducted on-site in different reference laboratories at Fundacao Oswaldo Cruz (Fiocruz), Rio de Janeiro, Brazil. The complete list of laboratory assays and sample types of each panel is provided on the study protocol [11].

### Clinical and laboratorial definitions

Leukopenia was defined as a white cell count <5000 cells/ μL. Thrombocytopenia was defined as a platelet <150.000/μL. Neutropenia was defined as neutrophils <1500 (10^9^ μL). Quick sequential organ failure assessment ≥ 2 was used to identify those with severe disease at high risk for poor outcomes.

Previous arboviruses exposure was evaluated using DPP^®^ ZDC IgM/IgG and IgM/IgG ELISA (Euroimmune, Germany) against ZIKV, DENV, and CHIKV. It is important to emphasize that we aimed to evaluate past/recent exposure with such assays and not necessarily correlate with the actual etiology of fever. Due to extensive serology cross-reaction that has been reported between DENV and ZIKV in regions where both flaviviruses co-circulated [15], participants with test results positive for both viruses (IgM and/or IgG) were classified as flaviviruses. Conversely, positive test results for IgM and/or IgG from only one of the viruses were classified accordingly (exposure to DENV only, ZIKV only, or CHIKV only). Co-infection was defined as having a serological response to more than one virus in a blood sample. Participants negative to all three studied arboviruses were classified as “No arbovirus”.

### Assessment of final diagnoses

Final diagnosis (in some cases, more than one) was retrospectively established based on all available clinical and laboratory data from the index and follow-visit, as was done by others (see Supplementary Table 1) [16]. These criteria were applied to each participant.

### Statistical analysis

Descriptive analysis was performed to characterize the distributions of several variables. A Chi-square test was used to compared categorical variables between study groups. Continuous variables were compared between the study groups using analysis of variance or Kruskal-Wallis test if those were found to be normally or non-normally distributed, respectively.

We dealt with missing data using the listwise deletion method (or complete case analysis).

For the clinical prediction model, we initially made descriptive univariate logistic regression analyses exploring each predictor and outcome’s crude association and selected variables with 2-sided *p* <0.020 on univariate analysis. Then, we conducted a forward stepwise logistic regression model selection, including only those predictors showing significance on a 10% significance level determined by a likelihood-ratio test. A 2-sided *p-value* less than 0.05 was considered statistically significant.

All statistical analyses were performed using SPSS software (version 26, IBM, Chicago, Illinois, USA) and Tableau Desktop (version 2020.4.2, Tableau, Seattle, Washington, USA).

## Results

### Enrollment and study sample characteristics

A total of 505 febrile participants were initially enrolled; five were subsequently excluded from the study, as shown in the study flowchart (Figure 2).

**Figure 2.**
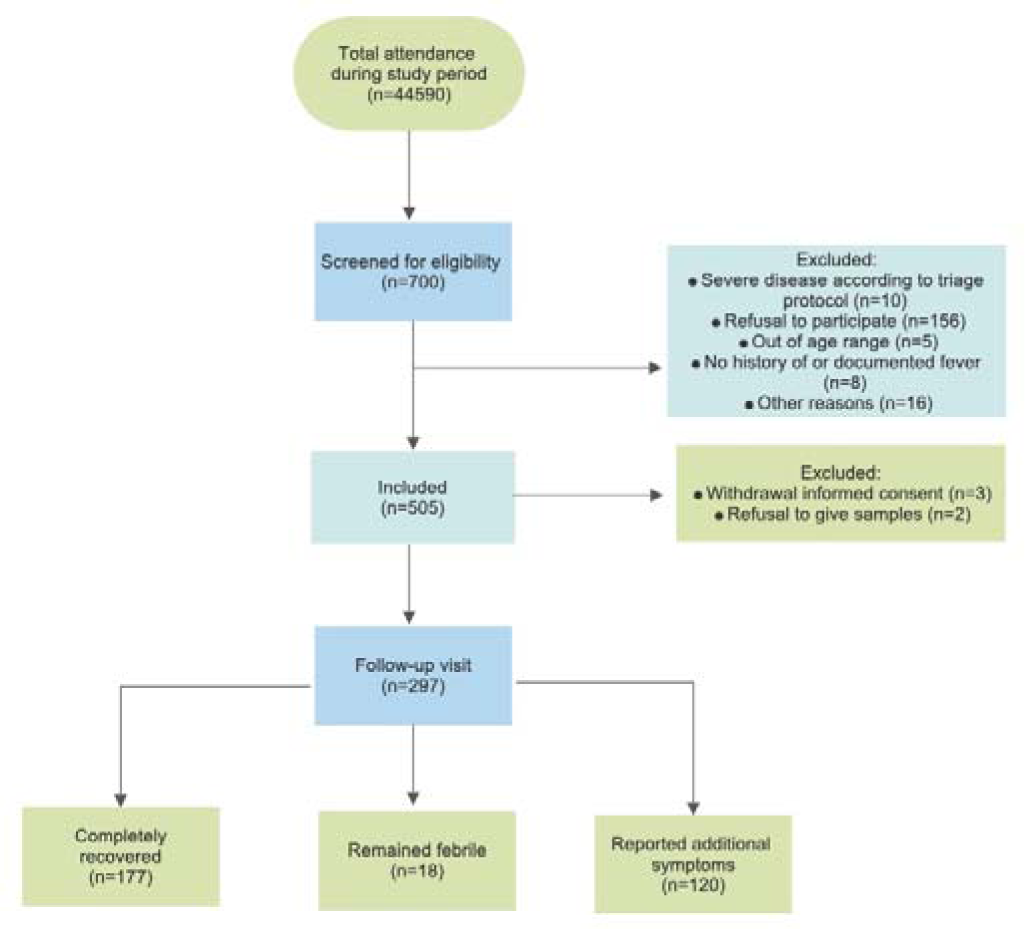
Study enrollment flow chart among participants with acute febrile illness, Rio de Janeiro, October 2018-July 2019.

Demographic, clinical, and laboratory characteristics of the 500-participants included in the study are described in Table 1. There was a balanced gender distribution (50.4% females), and the median duration of illness was 3 [2-4] days. Besides fever, the main complaints more frequently reported were headache (76.4%), arthralgia (54.4%), and cough (35.8%).

**Table 1.** Demographic, clinical and laboratory characteristics among non-severe febrile illness participants, Rio de Janeiro, Brazil, October 25, 2018 – July 31, 2019

| Characteristics | Total febrile population<br>(N=500) |
| --- | --- |
| **Demographic variables** |  |
| Female sex – no. (%) | 252 (50.4) |
| Age (years), median (IQR) | 26 (15-41) |
| Age (years) – no. (%) |  |
| ≤ 15 | 77 (15.4) |
| 16 – 26 | 85 (17) |
| 27 - 40 | 279 (55.8) |
| ≥ 41 | 59 (11.8) |
| **Clinical variables** |  |
| BMI (kg/m<sup>2</sup>), median (IQR) | 25.8 (20.1-29.8) |
| Mild upper arm circumference (cm) for those < 5 years, mean (±SD) | 176.4 (60.4) |
| Illness duration (days), median (IQR) | 3 (2-4) |
| **Symptoms** |  |
| Headache – no. (%) | 382 (76.4) |
| Photophobia – no. (%) | 137 (27.4) |
| Sore throat – no. (%) | 154 (30.8) |
| Rhinorrhea – no. (%) | 117 (23.4) |
| Cough – no. (%) | 179 (35.8) |
| Dysuria – no. (%) | 40 (8) |
| Diarrhea – no. (%) | 81 (16.2) |
| Joint pain – no. (%) | 272 (54.4) |
| **Medical past history** |  |
| Recent antibiotic use – no. (%) | 44 (8.8) |
| Comorbidities – no. (%) | 124 (24.8) |
| **Vital signs** |  |
| Temperature (°C), median (IQR) | 37.7 (36.7-38.4) |
| Heart rate (beats/min), mean (±SD) | 101 (21) |
| Respiratory rate (rate/min), median (IQR) | 21 (19-24) |
| Mean arterial pressure (mmhg), mean (±SD) | 83.5 (25.7) |
| **Laboratory variables** |  |
| WBC (cells/uL), median (IQR) | 7.280 (5.460-10.390) |
| Lymphocytopenia (10<sup>9</sup> uL) – no. (%) | 246 (49.5) |
| Platelets (uL), median (IQR) | 243 (196.000-297.000) |
| Creatinine (mg/dl), median (IQR) | 0.84 (0.68-1.07) |
| Total bilirubin (mg/dl), median (IQR) | 0.39 (0.26-0.56) |
| CRP > 5 mg/dl – no. (%) | 318 (63.9) |
| BMI stands for body mass index; CRP C-reactive protein; IQR interquartile range; SD standard deviation WBC white blood count |  |

### Laboratory investigation

As shown in Figure 3, clinicians requested the laboratory panel investigations as appropriate for each participant. None of the pathogens we tested for could be detected in 124 (24.8%) participants (Supplementary Table 2).

**Figure 3.**
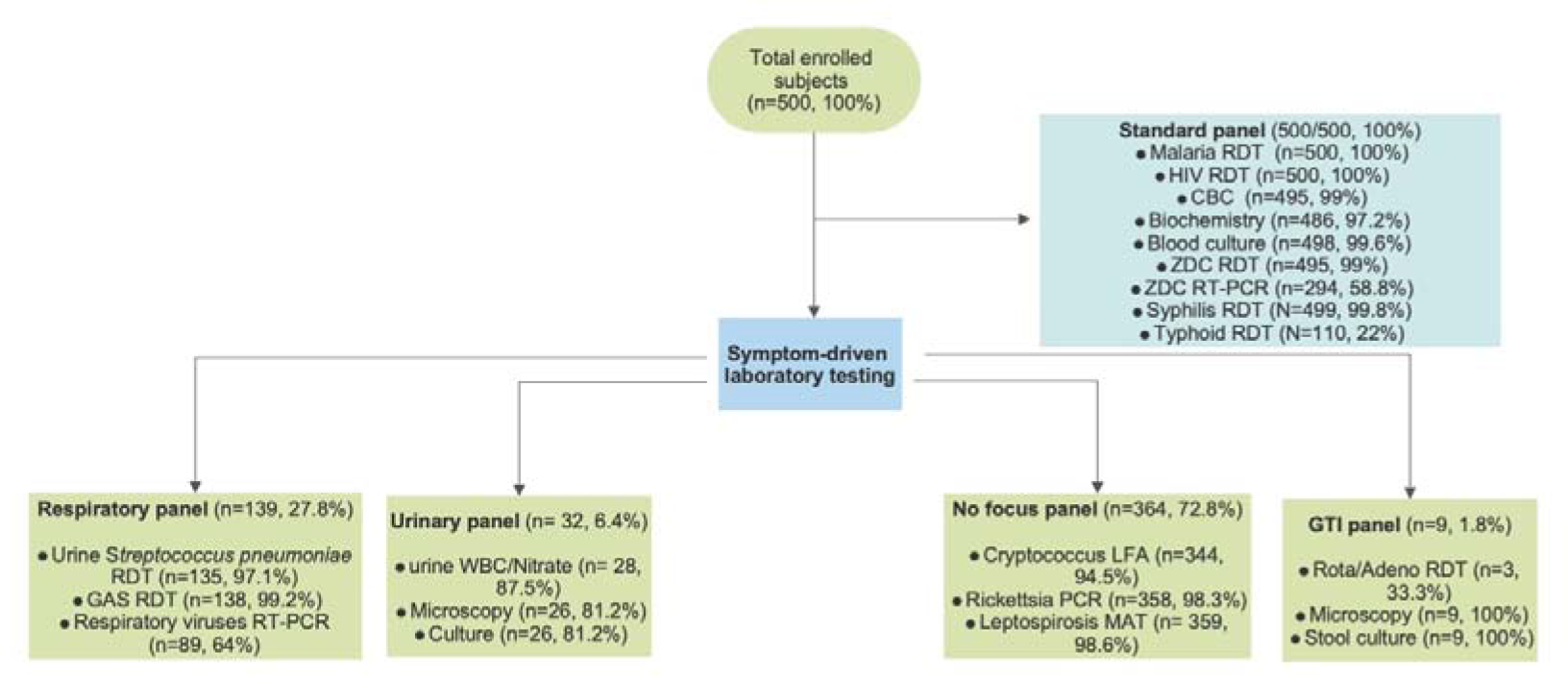
Flowchart of laboratory investigations conducted in acute febrile illness participants, Rio de Janeiro, October 2018-July 2019.

### Syndromic, clinical, and etiologic diagnosis

Overall, 824 distinct diagnoses were made (Figure 4). Systemic infection was the most frequent syndromic diagnosis (382/500, 76%), followed by acute respiratory tract infection (155/500, 31%) and urinary tract infection (23/500, 4.6%). Supplementary Figure 1 shows the distribution of the primary febrile syndromes by the week of illness onset. The diagnosis most prevalent for the participant with systemic infections were CHIKV (284/382, 74.3%), followed by Flaviviruses (214/382, 56%), and DENV (51/382, 13.3%). Viral upper respiratory tract infection (40/155, 25.8%), streptococcal tonsillitis (37/155, 23.8%), and viral pneumonia (10/155, 6.4%) accounted for the most prevalent diagnosis in those with an acute respiratory infection. *Escherichia coli* (10/23, 43.4%) was the predominant organism responsible for culture-confirmed urinary tract infection. None of the participants was positive for malaria.

**Figure 4.**
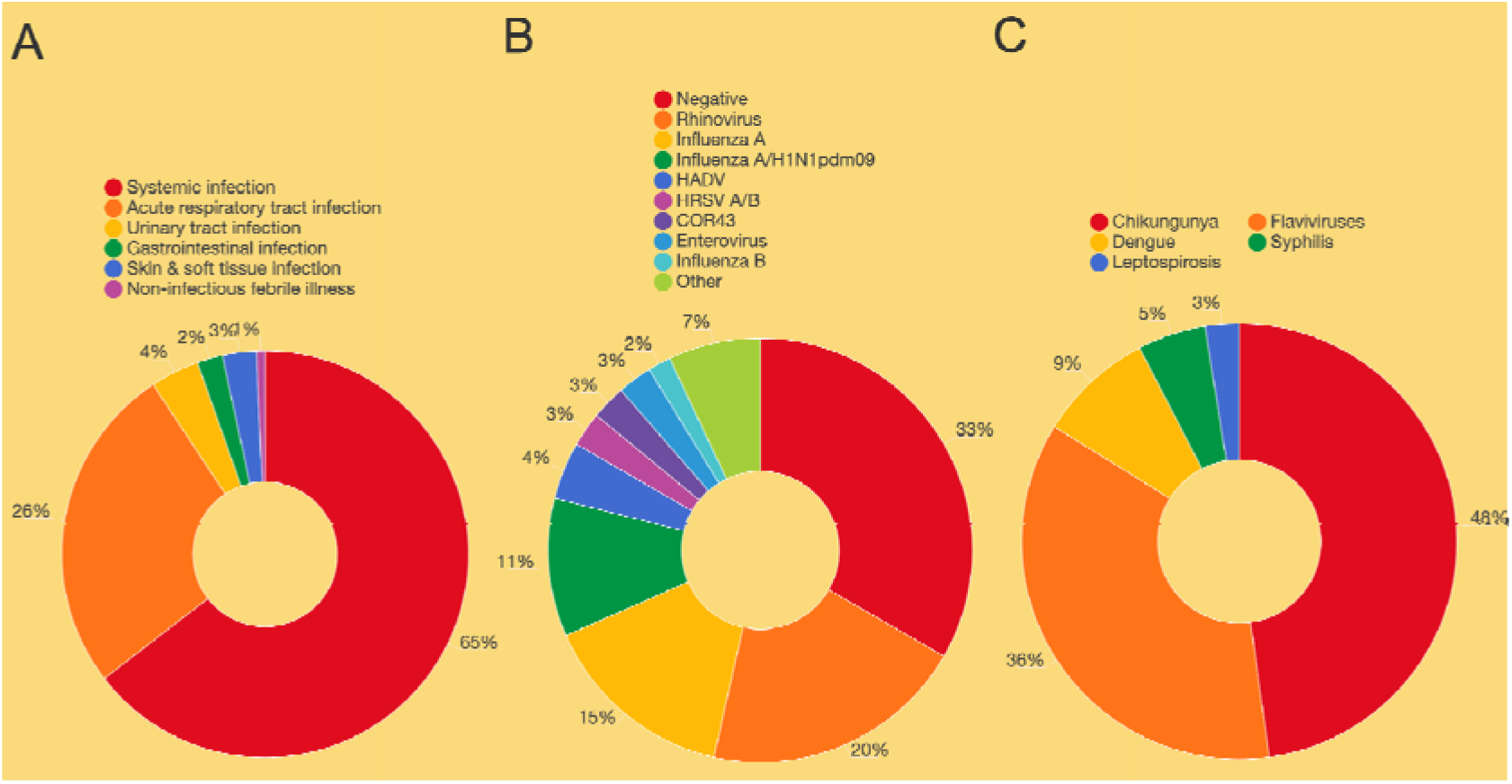
Distribution of 824 diagnoses among 500 non-severe febrile illness, Rio de Janeiro, Brazil, October 2018-July 2019 A. Febrile syndromes; B. Viral etiologies of the upper respiratory tract infection; and C. Microbial causes of systemic infection

The definitive etiologic diagnosis was made with a single pathogen in 139/500 (27.8%) participants, and two or more etiologic diagnoses were made in 249/500 (49.8%) participants.

Figure 5 shows the distribution of the etiologic diagnosis according to age group, respectively. Acute respiratory infection was predominantly diagnosed in the youngest participants (≤ 15 years in 44.6%); similar occurred for viral upper respiratory tract infection in this age group (45% of the total). Pyelonephritis was diagnosed most frequently in 19.5% of the young adults’ participants compared with 4.5% in the older adults. Confirmed CHIKV infection (i.e., by RT-PCR) was prevalent in all age groups, but the frequency increased with increasing age, accounting for 40% of all febrile participants ≤ 15 years-old versus 63.6% in participants ≥ 41 years-old.

**Figure 5.**
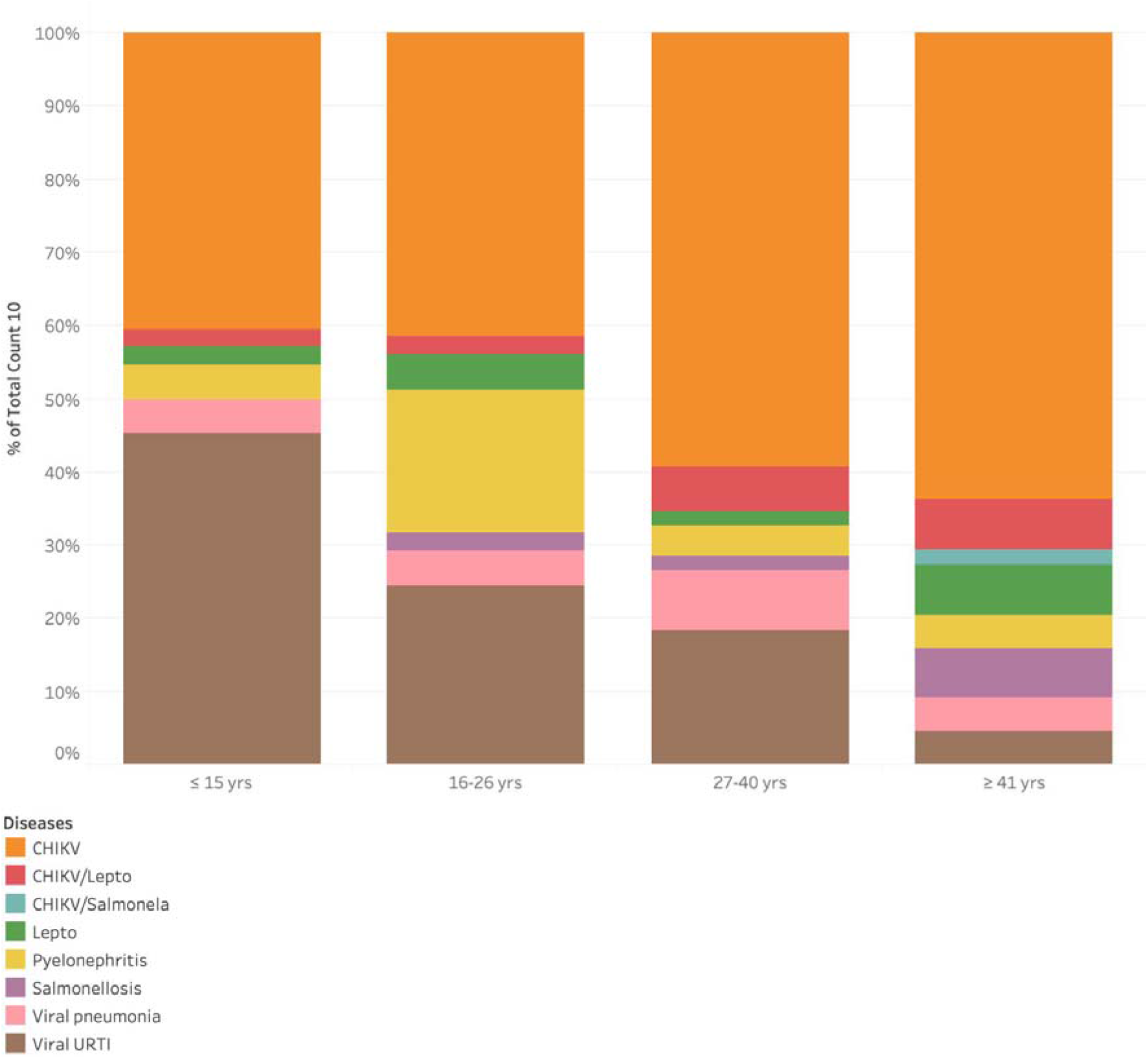
Distribution of the most common etiologic diagnosis according to age group, Rio de Janeiro, Brazil, October 2018-July 2019. CHIKV stands for chikungunya; Lepto leptospirosis; URTI upper respiratory tract infection

Supplementary Table 3 shows the main difference in demographic, clinical, and laboratory features according to the principal etiologic diagnosis.

### Previous arboviruses exposure

The DPP^®^ ZDC IgM/IgG results were available for 495 (99%) of the participants. DENV was confirmed in 369 (74.5%) participants, 128 (25.8%) alone and 31 (6.2%) associated with CHIKV (Figure 6). Flaviviruses accounted for 149 (30.1%). Antibodies against ZIKV were found in 218 (44%) participants, 3 (0.6%) alone, and 5 (1%) in association with antibodies against CHIKV. CHIKV was confirmed in 117 (23.6%) participants, 19 (3.8%) alone, and 36 (7.2%) in association with other arboviruses. Overall, 397 (80%) participants were positive for at least one arbovirus, and 80 (16%) were negative for all arboviruses tested. The ELISA results against the three arboviruses are shown in Supplementary Figure 2.

**Figure 6.**
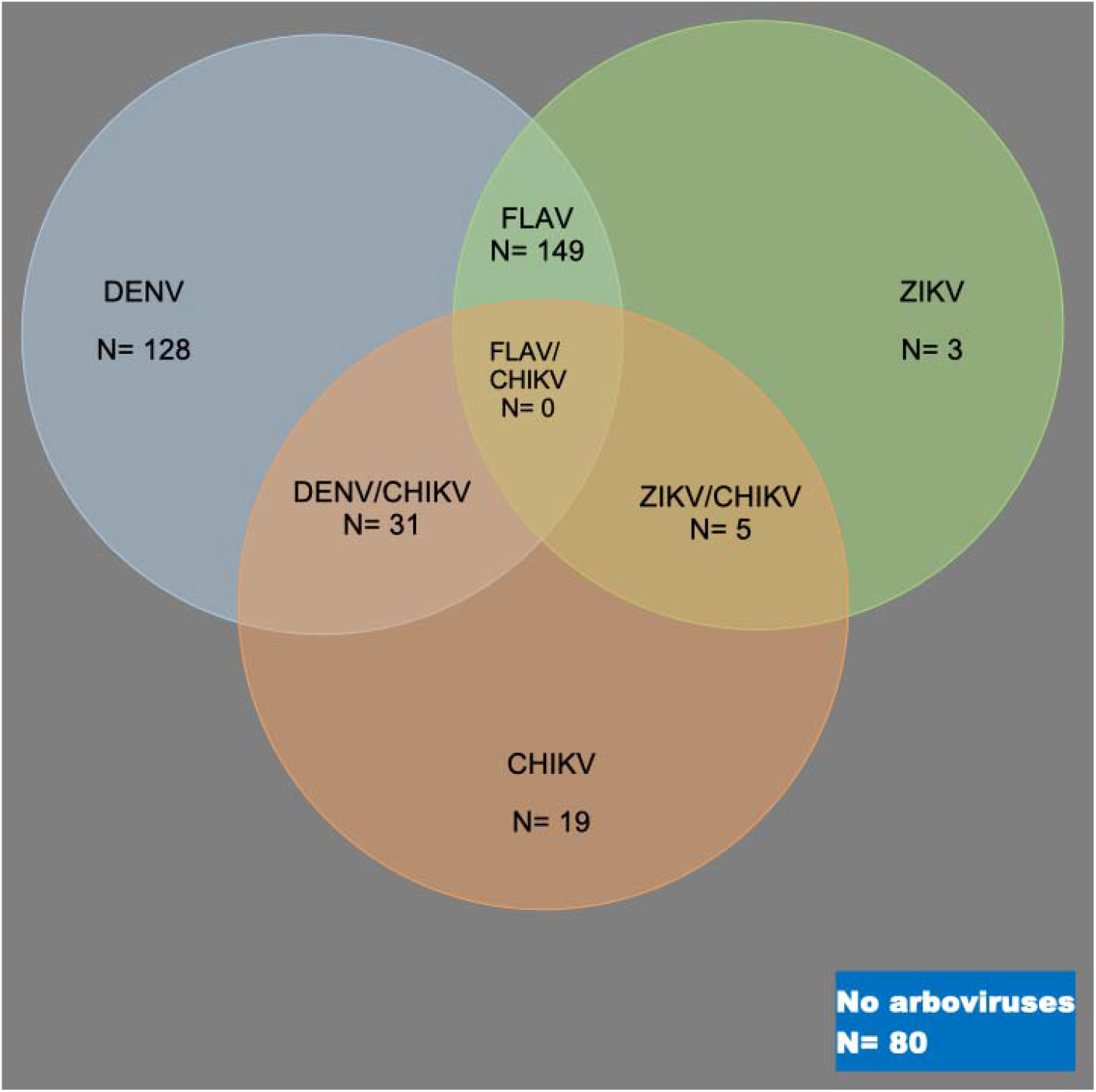
Venn diagram showing the distribution of the rapid diagnostic test arboviruses’ results in the sample cohort and the proportion of co-infection between the arboviruses, Rio de Janeiro, Brazil, October 2018-July 2019. DENV means dengue virus; CHIKV chikungunya; FLAV flaviviruses; ZIKV zika virus.

### RT-PCR-confirmed arboviruses infection

Two hundred ninety participants had samples collected for arbovirus RT-PCR assay. A hundred (100/290, 34%) participants were positive for CHIKV, and none of them were positive for the other two arboviruses.

### Clinical and laboratory presentation of CHIKV infection

Table 2 shows the main demographic, clinical, and laboratory characteristics of participants according to CHIKV (i.e., confirmed by RT-PCR). Chikungunya positive individuals were more likely to be male (63% *vs*. 46.4%), older (32.6 ± 16.2 *vs*. 25.8 ± 15.5 years), had a higher body mass index (27.6 [23.7-31.4] *vs*. 24.6 [18.8-29.4] kg/m^2^) and presented earlier (2 [1-3] *vs*. 3 [2-4] days) after symptom onset than their negative counterparts. Several symptoms were more prevalent in CHIKV positive patients compared with CHIKV negatives: headache (90% *vs*. 67.5%), redness of the eyes (38% *vs*. 12.9%), photophobia (38% *vs*. 18.6%), arthralgia (88% *vs*. 25.3%) and rash (54% *vs*. 6.7%), respectively.

**Table 2.**
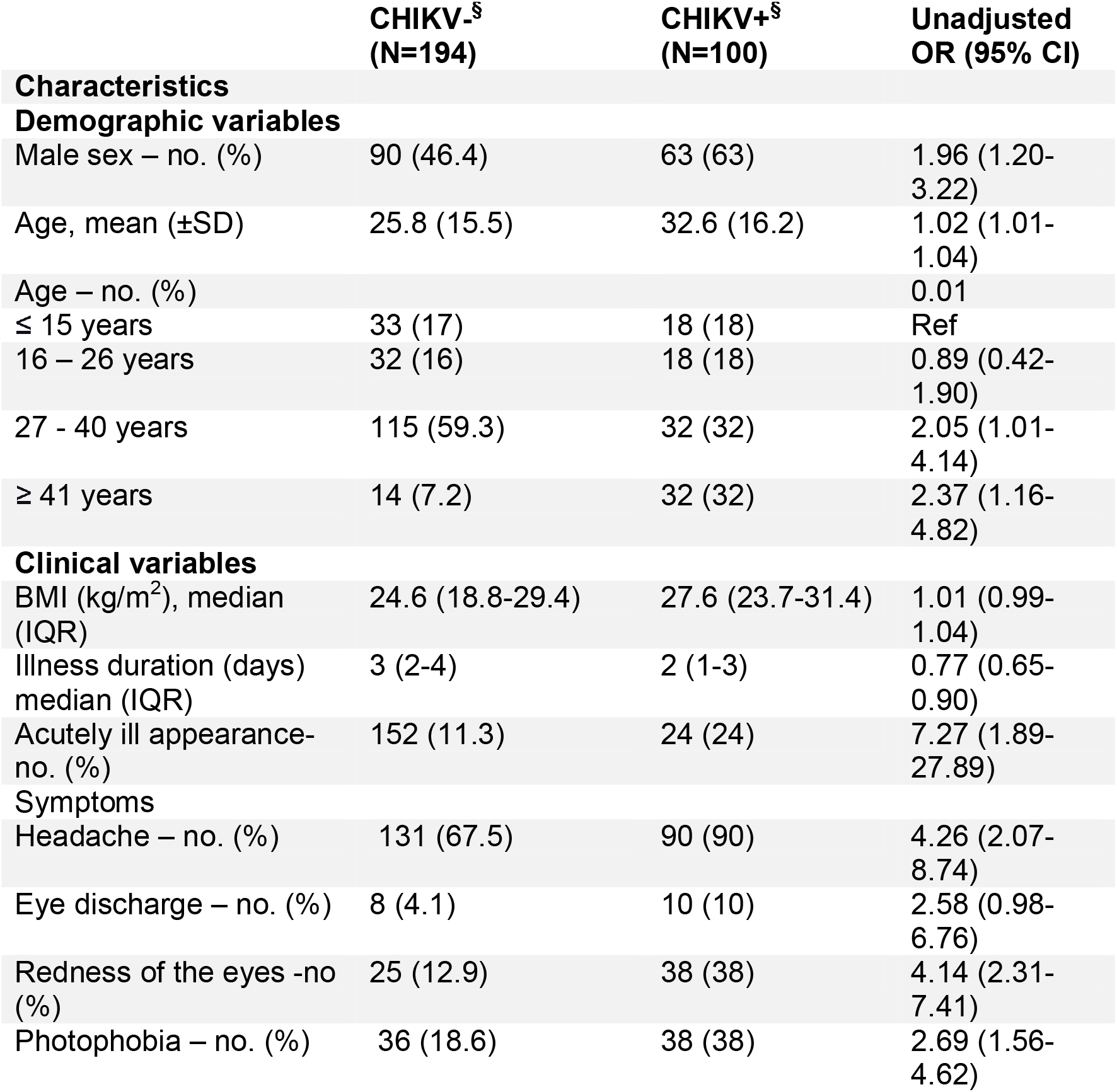
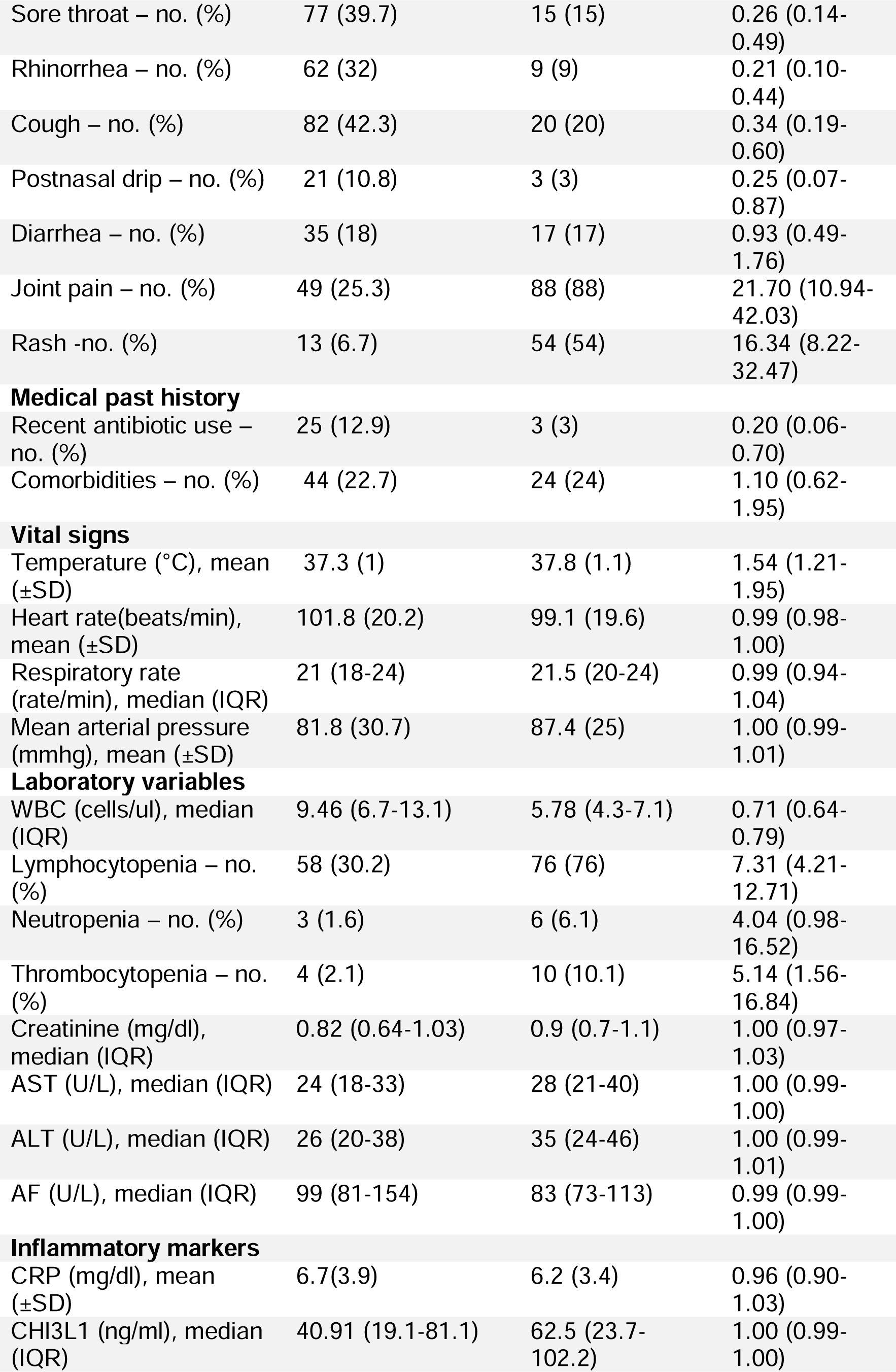
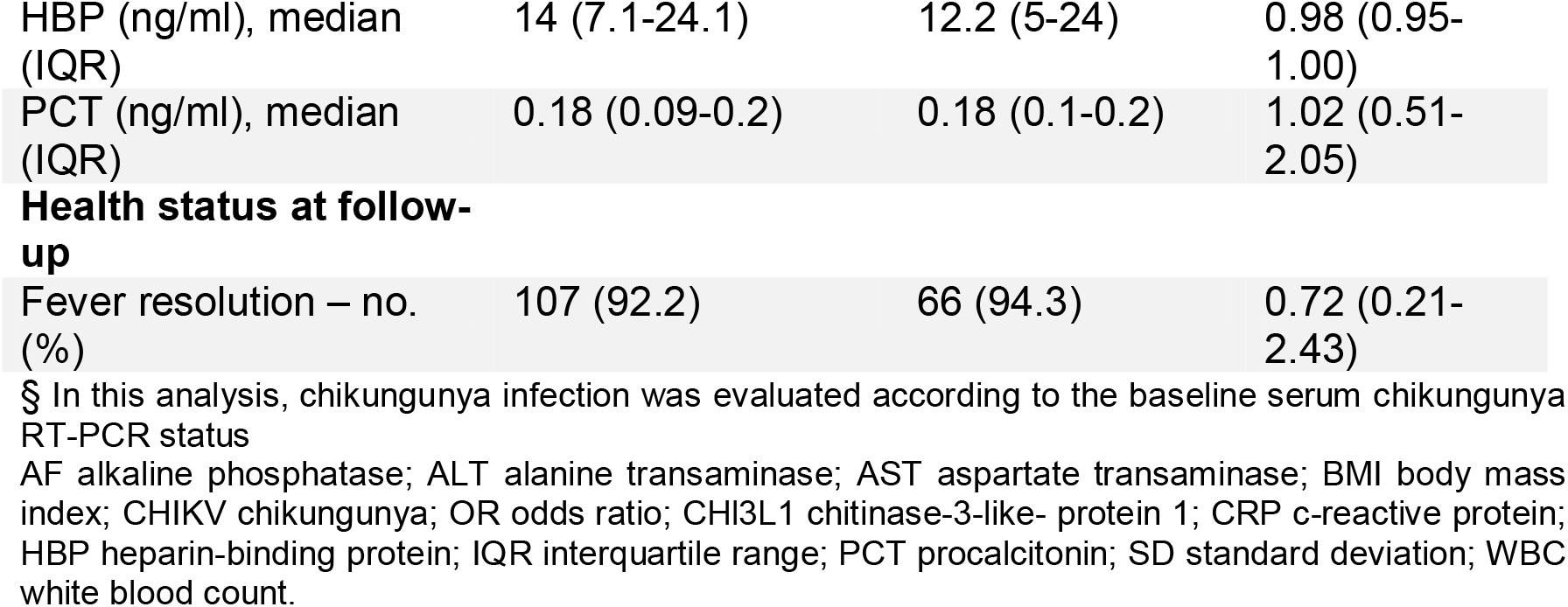
Demographic, clinical and laboratory characteristics of participants according to chikungunya infection, Rio de Janeiro, Brazil, October 25, 2018 – July 31, 2019

|  | CHIKV-<sup>s</sup><br>(N=194) | CHIKV+<sup>s</sup><br>(N=100) | Unadjusted<br>OR (95% CI) |
| --- | --- | --- | --- |
| **Characteristics** |  |  |  |
| **Demographic variables** |  |  |  |
| Male sex – no. (%) | 90 (46.4) | 63 (63) | 1.96 (1.20-3.22) |
| Age, mean (±SD) | 25.8 (15.5) | 32.6 (16.2) | 1.02 (1.01-1.04) |
| Age – no. (%) |  |  | 0.01 |
| ≤ 15 years | 33 (17) | 18 (18) | Ref |
| 16 – 26 years | 32 (16) | 18 (18) | 0.89 (0.42-1.90) |
| 27 - 40 years | 115 (59.3) | 32 (32) | 2.05 (1.01-4.14) |
| ≥ 41 years | 14 (7.2) | 32 (32) | 2.37 (1.16-4.82) |
| **Clinical variables** |  |  |  |
| BMI ($\text{kg/m}^2$), median (IQR) | 24.6 (18.8-29.4) | 27.6 (23.7-31.4) | 1.01 (0.99-1.04) |
| Illness duration (days) median (IQR) | 3 (2-4) | 2 (1-3) | 0.77 (0.65-0.90) |
| Acutely ill appearance- no. (%) | 152 (11.3) | 24 (24) | 7.27 (1.89-27.89) |
| **Symptoms** |  |  |  |
| Headache – no. (%) | 131 (67.5) | 90 (90) | 4.26 (2.07-8.74) |
| Eye discharge – no. (%) | 8 (4.1) | 10 (10) | 2.58 (0.98-6.76) |
| Redness of the eyes -no (%) | 25 (12.9) | 38 (38) | 4.14 (2.31-7.41) |
| Photophobia – no. (%) | 36 (18.6) | 38 (38) | 2.69 (1.56-4.62) |
| Sore throat – no. (%) | 77 (39.7) | 15 (15) | 0.26 (0.14-0.49) |
| Rhinorrhea – no. (%) | 62 (32) | 9 (9) | 0.21 (0.10-0.44) |
| Cough – no. (%) | 82 (42.3) | 20 (20) | 0.34 (0.19-0.60) |
| Postnasal drip – no. (%) | 21 (10.8) | 3 (3) | 0.25 (0.07-0.87) |
| Diarrhea – no. (%) | 35 (18) | 17 (17) | 0.93 (0.49-1.76) |
| Joint pain – no. (%) | 49 (25.3) | 88 (88) | 21.70 (10.94-42.03) |
| Rash -no. (%) | 13 (6.7) | 54 (54) | 16.34 (8.22-32.47) |
| **Medical past history** |  |  |  |
| Recent antibiotic use – no. (%) | 25 (12.9) | 3 (3) | 0.20 (0.06-0.70) |
| Comorbidities – no. (%) | 44 (22.7) | 24 (24) | 1.10 (0.62-1.95) |
| **Vital signs** |  |  |  |
| Temperature (°C), mean (±SD) | 37.3 (1) | 37.8 (1.1) | 1.54 (1.21-1.95) |
| Heart rate(beats/min), mean (±SD) | 101.8 (20.2) | 99.1 (19.6) | 0.99 (0.98-1.00) |
| Respiratory rate (rate/min), median (IQR) | 21 (18-24) | 21.5 (20-24) | 0.99 (0.94-1.04) |
| Mean arterial pressure (mmhg), mean (±SD) | 81.8 (30.7) | 87.4 (25) | 1.00 (0.99-1.01) |
| **Laboratory variables** |  |  |  |
| WBC (cells/ul), median (IQR) | 9.46 (6.7-13.1) | 5.78 (4.3-7.1) | 0.71 (0.64-0.79) |
| Lymphocytopenia – no. (%) | 58 (30.2) | 76 (76) | 7.31 (4.21-12.71) |
| Neutropenia – no. (%) | 3 (1.6) | 6 (6.1) | 4.04 (0.98-16.52) |
| Thrombocytopenia – no. (%) | 4 (2.1) | 10 (10.1) | 5.14 (1.56-16.84) |
| Creatinine (mg/dl), median (IQR) | 0.82 (0.64-1.03) | 0.9 (0.7-1.1) | 1.00 (0.97-1.03) |
| AST (U/L), median (IQR) | 24 (18-33) | 28 (21-40) | 1.00 (0.99-1.00) |
| ALT (U/L), median (IQR) | 26 (20-38) | 35 (24-46) | 1.00 (0.99-1.01) |
| AF (U/L), median (IQR) | 99 (81-154) | 83 (73-113) | 0.99 (0.99-1.00) |
| **Inflammatory markers** |  |  |  |
| CRP (mg/dl), mean (±SD) | 6.7(3.9) | 6.2 (3.4) | 0.96 (0.90-1.03) |
| CHI3L1 (ng/ml), median (IQR) | 40.91 (19.1-81.1) | 62.5 (23.7-102.2) | 1.00 (0.99-1.00) |
| HBP (ng/ml), median (IQR) | 14 (7.1-24.1) | 12.2 (5-24) | 0.98 (0.95-1.00) |
| PCT (ng/ml), median (IQR) | 0.18 (0.09-0.2) | 0.18 (0.1-0.2) | 1.02 (0.51-2.05) |
| **Health status at follow-up** |  |  |  |
| Fever resolution – no. (%) | 107 (92.2) | 66 (94.3) | 0.72 (0.21-2.43) |

§ In this analysis, chikungunya infection was evaluated according to the baseline serum chikungunya RT-PCR status
AF alkaline phosphatase; ALT alanine transaminase; AST aspartate transaminase; BMI body mass index; CHIKV chikungunya; OR odds ratio; CHI3L1 chitinase-3-like- protein 1; CRP c-reactive protein; HBP heparin-binding protein; IQR interquartile range; PCT procalcitonin; SD standard deviation; WBC white blood count.

Most laboratory parameters were remarkably different at the index visit. We found a positive correlation between CHIKV viral load and serum lymphocyte count (Pearson coefficient=0.46, *p*=0.01).

Children (24/100, 24%) infected with CHIKV more frequently reported (70.8% *vs*. 48.7%), and arthralgia were less present (66.7% *vs*. 94.7%) compared with the adult’s counterpart (76/100, 76%). Figures 7 shows clinical photographs from participants positive for CHIKV in our cohort (These photographs contain sensible participants information that is why, we advise readers to contact the corresponding author in case they request access to these materials).

We then investigated the factors independently associated with positive RT-PCR CHIKV at the index visit. We found that the absence of cough [(aOR: 0.30 (0.13-0.69)], arthralgia [(aOR: 11.87 (5.35-26.32)], rash [(aOR: 7.07 (3.06-16.30)], temperature [(aOR: 1.67 for each point increase in temperature, 95% CI: 1.16-2.40)], and leucopenia [(aOR: 3.57 (1.24-10.28)] were associated with CHIKV (Table 3).

**Table 3.**
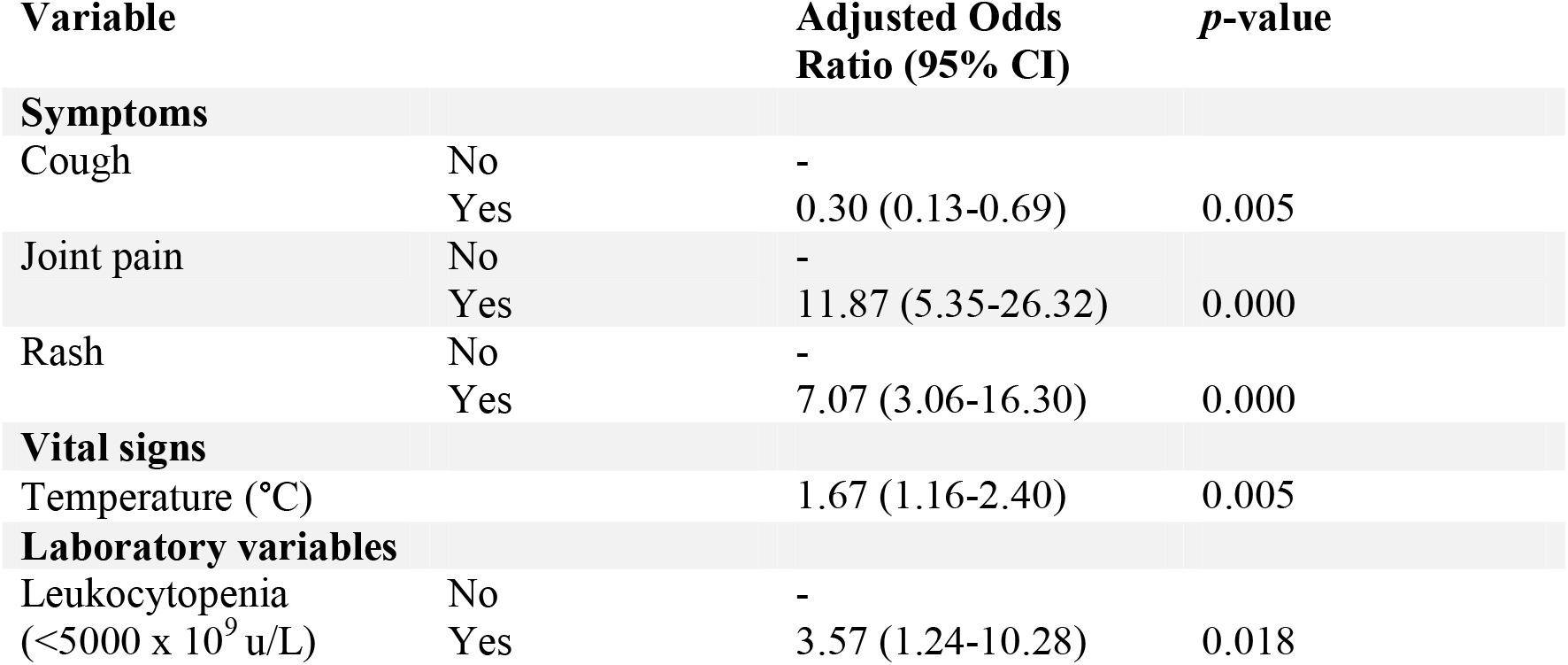
Prediction model for 294 acute febrile illness patients evaluated for Chikungunya infection, Rio de Janeiro, Brazil, October 25, 2018 – July 31, 2019.

Analysis of the temporal trends demonstrated that CHIKV occurred in 2018 and continued through 2019, in nearly the total study period [Supplementary Figure 3].

### Outcomes

A total of 297/500 (59.4%) patients completed the follow-visit (Fig 2) in a median of 16.5 (±5.5) days. Of those who returned for the next visit, 18 (6.1%) had fever relapse, while 279 (93.9%) reported fever resolution after a mean of 2.2 (±2) days after the index visit.

New or persistent symptoms occurred in 120/297 (40%) patients. The most common symptom reported was osteomuscular (45%), followed by respiratory (17.5%), miscellaneous (17.5%), and gastrointestinal (16.6%). Of the 297 participants that attended the follow-visit, 70/297 (23.5%) and 116/297 (39%) were classified as CHIKV positive (i.e., presumptive or confirmatory infection) and negative at index visit, respectively. CHIKV positive patients were more likely to experience persistent osteomuscular symptoms than CHIKV negative [OR: 10.18 (3.64-28.45)].

## Discussion

The syndrome of fever is a common presenting complaint among persons seeking healthcare in resource-constrained settings, yet the global health community has not approached it comprehensively [17]. Health care workers working in low-resource settings mostly rely on history taking and physical examination to determine the focus (or the absence of it) and the probable cause of infection, of which acute respiratory infection, gastroenteritis, and urinary infection are the most prevalent syndromes reported [16]. However, the non-specific clinical presentation of many infections that cause fever makes it difficult to distinguish one from another base on history alone. Here, using a standardized and harmonized syndromic approach with an extensive set of clinical and laboratory data, we were able to identify the possible cause of fever (confirmed or suspected) in 75% of the non-severe participants seeking care at the primary level in Rio de Janeiro, Brazil. Systemic infection was the most frequent syndromic diagnosis (76%), followed by acute respiratory tract infection (31%) and urinary tract infection (5%). Evidence of a viral process was implicated in the majority of them (80%), and the diagnoses most prevalent were CHIKV (57%), followed by Flaviviruses (43%), respiratory viruses (10%), and DENV (8%). Next, our findings suggest that a set of clinical and laboratory markers - the absence of cough, the presence of arthralgia, rash, higher temperature, and leucopenia - could predict those more likely to be laboratory-confirmed CHIKV amidst an CHIVK epidemic, highlighting the importance of considering these factors when developing prediction rules to aid in the clinical management of arboviruses suspected patients. Last, our data suggest that laboratory-confirmed CHIKV participants were more likely to report persistent arthritis in weeks after illness onset, which agrees with other data [18].

Our study has several limitations. First, considering that we conducted recruitment in 10-months in study sites representative of urban Rio de Janeiro, and when a large epidemic of CHIKV was ongoing, this means that the generalizability of our findings is uncertain. Due to the outbreak, the CHIKV detection rate was overestimated as a cause of fever in the study cohort and accounted for more than half of all fever cases. Second, our findings indicate that in our setting a high percentage of the febrile systemic syndrome is likely due to a Flavivirus. Discriminating the contribution of either DENV or ZIKV without performing a plaque neutralization assay is however challenging. Thus, we chose to categorize this uncertainty as Flaviviruses. Third, the higher proportion of arboviruses found in our study is at risk of being wrongly interpreted as acute infection instead of previous/recent infection. Sparse data suggest that specific serum antibodies for ZIKV, DENV, and CHIKV might be detectable for months or perhaps years [19]. Fourth, diagnostic tests deployed as part of the biomarker evaluation study [11] did not represent the gold standard of testing for etiologies and hence causative agents might have been missed or misclassified [20]. This might impact the pathogen detection rate observed in our cohort, and also the quality of data. Fifth, for logistical reasons we were unable to collect some samples (i.e., nasal swab, urine, stool) and thus conduct the corresponding diagnostic assays in a proportion of participants, mainly those who were enrolled in emergency departments. This resulted in incomplete diagnostic information, which means that we had to extrapolate prevalence from the tested population to the untested population, potentially introducing bias. Sixth, follow-up was not possible for 40% of the participants, which means outcome data are incomplete, and we might have misinterpreted the proportion of participants that completely recovered at follow-up. Several strategies were used to reduce the rate of follow-up failure: phone calls, text messages, and social media contacts, but with limited success. Seventh, we did not consider febrile participants with a different spectrum of severity, underrecognizing essential pathogens known to occur in our region that were not detected. For instance, bloodstream infections due to bacteria contributed merely 0.6% of the total causes of non-severe fevers in outpatients, which is in striking contrast with 9.8% found in a study that enrolled severe febrile inpatient in Northern Tanzania [21]. Finally, inclusion of a suitable control group would have allowed the calculation of attributable fractions for individual pathogens, which should be considered for future febrile illness research.

Our findings strongly suggest that arboviruses play a considerable role in explaining the regions’ AFI burden, highlighted by the fact that 80% of the participants had a serologic response to any of the three arboviruses tested. This fact corroborates with a survey conducted in Rio, which found that more than 80% of the city’s residents have had a previous contact with dengue, zika or chikungunya [22].

In urban Rio de Janeiro, most (75%) of the febrile non-severe participants were identified with a fever etiology, and the remaining ones were left without a definitive diagnosis even after extensive testing. This is high compared to other studies that only identify up to 41% [23]. The majority of febrile illnesses were attributed to viral pathogens that typically do not require antibiotic treatment. Unsurprisingly, CHIKV was the leading cause of AFI, albeit DENV might have been substantially underestimated as mixed DENV and ZIKV infections were assumed as Flaviviruses. Furthermore, we demonstrate that CHIKV affects all age groups, predominantly older adults who may be at higher risk of developing medical complications and death. Finally, there is an urgent need to strengthen the local laboratory capacity with affordable fever diagnostics to stem the ever-increasing problem of antimicrobial resistance and, ultimately, improve the clinical management algorithms of outpatients in low-resource setting.

## Data Availability

All data produced in the present work are contained in the manuscript.

## Contributions

All authors critically reviewed the manuscript, gave their final approvals, and are accountable for accuracy and integrity.

## Acknowledgments

The authors want to thank all participants and their family members who have contributed to these efforts. The authors also want to acknowledge the help of all of the clinical, laboratory, and administrative staff at the UPA Manguinhos, UPA Rocha Miranda, Centro de Saude Escola Germano Sinval Faria (CSEGSF) and Clinica da Familia Armando Palhares Aguinaga (CFAPA). This study would not have been possible without their support. JM received a scholarship from Fundação de Amparo a Pesquisa do Estado do Rio de Janeiro (FAPERJ) for his PhD study in Brazil.

## REFERENCES

1. Ferreira UM, Castro MC. Malaria situation in Latin America and the Caribbean: residual and resurgent transmission and challenges for control and elimination. Methods Mol Biol. 2019; 2013:57–70.

2. Moreira J, Barros J, Lapouble O et al. When fever is not malaria in Latin America: a systematic review. BMC Med. 2020 18(1):294.

3. World Health Organization IMAI district clinician manual: hospital care for adolescents and adults: guidelines for the management of illness with limited resources. 2011. Geneva. World Health Organization

4. World Health Organization Pocket book of hospital care for children: guidelines for the management of common illness with limited resources. 2005. Geneva. World Health Organization.

5. Reyburn H. Mbatia R, Drakeley C, et al. Overdiagnosis of malaria in patients with severe febrile illness in Tanzania: a prospective study. BMJ. 2004; 329: 1212–15.

6. Moreira J, Bressan CS, Brasil P, Siqueira AM. Epidemiology of acute febrile illness in Latin America. Clin Microbiol Infect. 2018 24(8): 827–35.

7. Schatzmayr HG, Nogueira RM, Travassos da Rosa AP. An outbreak of dengue virus at Rio de Janeiro –1986. Mem Inst Oswaldo Cruz. 1986 81(2): 245–6.

8. Nunes MR, Faria NR, de Vasconcelos JM et al. Emergence and potential for spread of chikungunya in Brazil. BMC Med. 2015 13:102.

9. Zanluca C, de Melo VCA, Mosimann ALP et al. First report of autochthonous transmission of Zika virus in Brazil. Mem Inst Oswaldo Cruz. 2015 110 (4): 569–72.

10. Boletim Epidemiologico Arboviroses Nº 002/2019. Cenario Epidemiologico: Dengue, Chikungunya e Zika no estado RJ. 1º Semetre de 2019. Secretaria de Estado de Saude de Saude do Rio de Janeiro. Http://www.riocomsaude.rj.gov.br/Publico/MostrarArquivo.aspx?C=F%2BJ77ZiVqng%3D Accessed on March 10th 2021.

11. Escadafal C, Geis S, Siqueira AM, et al. Bacterial versus non-bacterial infections: a methodology to support use-case-driven product development of diagnostics. BMJ Glob Health. 2020 5(10)e:003141.

12. Barcellos C. Zaluar A. [Homicides and territorial struglles in Rio de Janeiro favelas]. Rev Saude Publica. 2014 48(1): 94–102.

13. Lanciotti RS, Calisher CH, Gubler DJ, Chang GJ, Vorndam AV. Rapid detection and typing of dengue viruses from clinical samples by using reverse transcriptase-polymerase chain reaction. J Clin Microbiol. 1992 30:545–51.

14. Namekar M, Ellis EM, O’Connell M, et al. Evaluation of a new commercially available immunoglobulin M capture enzyme-linked immunosorbent assay for diagnosis of dengue virus infection. J Clin Microbiol. 2013 51(9):3102–6.

15. Feliz AC, Souza NCS, Figueiredo WM et al. Cross reactivity of commercial anti-dengue immunoassays in patients with acute Zika virus infection. J Med Virol. 2017 89(8): 1477–79.

16. D’Acremont V, Kilowoko M, Kyungu E, et al. Beyond malaria – causes of fever in outpatient Tanzanian children. N Engl J Med. 2014 370 (9):809–17.

17. Crump JA. Time for a comprehensive approach to the syndrome of fever in the tropics. Trans R Soc Trop Med Hyg. 2014 108(2): 61–62.

18. Feldstein LR, Rowhani-Rahbat A, Stales JE, Weaver MR, Halloran ME, Ellis EM. Persistent arthralgia associated with Chikungunya virus outbreak, US Virgin Islands, December 2014-February 2016. Emerg Infect Dis. 2017 23(4): 673–6.

19. Bozza FA, Moreira-Soto A, Rockstroh A, et al. Diffeential shedding and antibody kinetics of zika and chikungunya viruses, Brazil. Emerg Infect Dis. 2019 25(2):311–315.

20. Olsen SJ, Pruckler J, Bibb W, et al. Evaluation of rapid diagnostic tests for typhoid fever. J Clin Microbiol. 2004 42: 1885–89.

21. Crump JA, Morrissey AB, Nicholson WL, et al. Etiology of severe non-malaria febrile illness in Northern Tanzania: a prospective cohort study. PLoS Negl Trop Dis. 2013 7(7): e2324.

22. Perisse ARS, Sousa-Santos R, Duarte R, et al. Zika, dengue and chikungunya population prevalence in Rio de Janeiro city, Brazil, and the importance of serovalence studies to estimate the real number of infected individuals. PLoS One. 2020 15(12): e0243239.

23. Mayxay M, Castonguay-Vanier J, Chansamouth V, et al. Causes of non-malarial fever in Laos: a prospective study. Lancet Glob Health. 2013 1(1): e46–54.

