## Supplementary Information for "Addressing Acute Febrile Illness Using a Syndromic Approach During A Chikungunya Epidemic in Rio de Janeiro, Brazil: A Prospective Observational Study"

**Appendix Table of Contents**

This supplementary material has been provided by the authors to give readers additional information about their work.

**Supplementary Tables**

- Supplementary Table 1. Summary of the clinical and laboratory criteria to establish definitive diagnosis, Rio de Janeiro, Brazil, October 2018-July 2019
- Supplementary Table 2. Number and proportion of enrolled participants with acute febrile illness by pathogens detected, Rio de Janeiro, Brazil, October 25, 2018 – July 31, 2019
- Supplementary Table 3. Demographic, clinical and laboratory characteristics of study participants by the man etiologic diagnosis, Rio de Janeiro, Brazil, October 25, 2018 – July 31, 2019
- **Supplementary Figures**
- Supplementary Figure 1. Distribution of the main febrile syndromes in participants with acute febrile illness by week of illness onset, Rio de Janeiro, October 2018-July 2019
- Supplementary Figure 2. Venn diagram showing the distribution of the ELISA arboviruses’ results in the sample cohort and the proportion of co-infection between the arboviruses, Rio de Janeiro, Brazil, October 2018-July 2019
- Supplementary Figure 3. Distribution of participants tested for Chikungunya RT-PCR according to week of illness onset, Rio de Janeiro, Brazil, October 2018-July 2019

**Assessment of final diagnoses**

Final diagnosis (in some cases, more than one) was retrospectively established based on all available clinical and laboratory data from the index and follow-visit, as was done by previous studies. These criteria were applied to each participant.

Supplementary Table 1. Summary of the clinical and laboratory criteria to establish definitive diagnosis, Rio de Janeiro, Brazil, October 2018-July 2019

| Syndromic diagnosis | Clinical presentations considered for the diagnosis | Additional criteria needed for a documented diagnosis (usually laboratory test or X-ray) |
| --- | --- | --- |
| Malaria | | |
| Malaria | - All presentations | - Positive malaria RDT or blood smear |
| Typhoid | | |
| Typhoid | - All presentations | - Positive typhoid RDT OR blood or stool culture positive for *Salmonella typhi* |
| Meningitis | | |
| Meningitis | - Neck stiffness OR bulging fontanel OR Kerning/Brudzinski OR convulsions OR abnormal consciousness | - CSF culture positive for any microorganism potentially involved in meningitis other than typical contaminants - Positive Crag test |
| Urinary tract infection | | |
| Urinary tract infection (i.e., pyelonephritis, cystitis) | - Dysuria, hematuria or polyuria | - Positive urine culture (non-contaminant bacteria with bacteria 10^4^ CFU/ml, other than mixed growth) |
| Skin, mucosal and osseous purulent infections | | |
| Significant skin infection (i.e., furunculosis, empyema, skin abscess, cellulitis, erysipelas) | - Pain, local swelling, local tenderness, refusal to move the affected limb/arm, visible localized wound - Pus discharge | - Positive culture of pus or collected liquid |
| Gastroenteritis | | |
| Amoebic gastroenteritis | - Diarrhea ≥ 3 watery stools/day | - Stool direct examination positive for Amoeba |
| Bacterial gastroenteritis |  | - Stool culture positive for enterotoxic *Escherichia coli*, Salmonella or *Shigella spp*. |
| Gastroenteritis of unknown origin |  | - No Amoeba or bacteria on microscopy and culture |
| Viral gastroenteritis |  | - Positive rota- or adenovirus RDT |
| Acute respiratory tract infections | | |
| Upper respiratory tract infection  (rhinitis, rhinosinusitis, pharyngitis, tonsillitis, acute otitis media) | - Rhinitis: cough, runny nose/Nasal discharge, mouth breathing - Pharyngitis: cough, throat pain, difficulty in swallowing/ drooling of saliva. Pharyngeal erythema/inflammation - Tonsillitis: throat pain OR pharyngeal redness AND enlarged tonsils. We subclassified tonsillitis into streptococcal and non-streptococcal aetiology according to GAS RDT results - Acute otitis media: history of ear pain, auricular discharge OR bulging tympanum OR erythematous tympanum | - Nasopharyngeal swab positive for viruses - GAS RDT positive |
| Lower acute respiratory tract infection  (bronchitis, bronchiolitis,  pneumonia) | - Bronchitis/bronchiolitis: cough or difficult breathing with abnormal breathing rate and abnormal chest auscultation (wheezing, rhonchi). - Non-radiological pneumonia: Cough AND difficult breathing with fast breathing OR chest indrawing/nasal flaring/grunting OR dyspnoea OR abnormal chest auscultation (decreased or bronchial breath sounds, crackles, abnormal vocal resonance, pleural rub). No “primary end-point consolidation” on CXR OR CXR not interpretable OR CXR not done   Radiological pneumonia: Cough AND difficult breathing with fast breathing OR chest indrawing/nasal flaring/grunting OR dyspnoea OR abnormal chest auscultation (decreased or bronchial breath sounds, crackles, abnormal vocal resonance, pleural rub). “Primary end-point consolidation” on CXR visible. | - “Bronchitis” on CXR - PCR positive for one or more virus(es) on nasopharyngeal swabs - Positive *Streptococcus pneumoniae* antigen RDT on urine - Chest X-ray showing conscript consolidation, lobular infiltrates or heterogeneous opacities |
| Non-infectious febrile illness | | |
| Non-infectious febrile illness | - All presentations | - No bacterial, viral or malaria parasite isolated, no specific positive tests OR laboratory tests results (during this study) do not fit with clinical picture and evolution |
| Systemic infection | | |
| Bacteremia | - All presentations | - Blood culture for bacteria other than typical contaminants (i.e., Coagulase negative-staphylococcus, *Bacillus spp*., viridans group streptococci, *Corynebacterium spp*., *Propionibacterium spp*., *Micrococcus spp*.) |
| Flaviviruses | - All presentations | - Concurrent antibodies for DENV ELISA and ZIKV ELISA |
| Dengue | - All presentations | - Presumptive dengue infection: positive DENV ELISA IgM and/or IgG assay **without** a reactivity for ELISA ZIKV IgM and/or IgG |
|  |  | - Confirmed dengue infection: positive dengue PCR |
| Zika | - All presentations | - Presumptive zika infection: positive zika ELISA IgM and/or IgG assay **without** having a reactivity for DENV ELISA IgM and/or IgG |
|  |  | - Confirmed zika infection: positive zika PCR |
| Chikungunya | - All presentations | - Presumptive chikungunya infection: positive chikungunya IgM assay |
|  |  | - Confirmed chikungunya infection: positive chikungunya PCR |
| Leptospirosis | - All presentations | - Positive microscopic agglutination test for *Leptospira spp*. |
| Syphilis | - All presentations | - Positive syphilis RDT |
| Rickettsiosis | - All presentations | - Positive PCR for Rickettsia |

Supplementary Table 2. Number and proportion of enrolled participants with acute febrile illness by pathogens detected, Rio de Janeiro, Brazil, October 25, 2018 – July 31, 2019

| Pathogen | Specimen tested | Method of detection | Number of participant with positive detection / total number of participants tested (n/N) |
| --- | --- | --- | --- |
|  |  |  | Total (n=500) |
| Viruses |  |  |  |
| Positive for at least 1 respiratory virus^¶^ | NP | RT-PCR | 51/89 (57.3%) |
| Rhinovirus | NP | RT-PCR | 23/89 (25.8%) |
| Influenza A | NP | RT-PCR | 17/89 (19.1%) |
| Influenza A/H1N1pdm09 | NP | RT-PCR | 12/89 (13.4%) |
| HADV | NP | RT-PCR | 5/89 (5.6%) |
| HRSV A/B | NP | RT-PCR | 3/89 (3.3%) |
| COR43 | NP | RT-PCR | 3/89 (3.3%) |
| Enterovirus | NP | RT-PCR | 3/89 (3.3%) |
| Influenza B | NP | RT-PCR | 2/89 (2.2%) |
| HPIV1 | NP | RT-PCR | 2/89 (2.2%) |
| HPIV4 | NP | RT-PCR | 2/89 (2.2%) |
| HBoV | NP | RT-PCR | 2/89 (2.2%) |
| COR229 | NP | RT-PCR | 1/89 (1.1%) |
| COR63 | NP | RT-PCR | 1/89 (1.1%) |
| HKU | NP | RT-PCR | 0/89 (0%) |
| HPIV2 | NP | RT-PCR | 0/89 (0%) |
| HPIV3 | NP | RT-PCR | 0/89 (0%) |
| HMPV A/B | NP | RT-PCR | 0/89 (0%) |
| HPeV | NP | RT-PCR | 0/89 (0%) |
| Adenovirus | Stool | Antigen detection | 0/3 (0%) |
| Rotavirus | Stool | Antigen detection | 0/3 (0%) |
| Chikungunya | Blood | RT-PCR | 100/294 (34%) |
| Chikungunya | Blood | ELISA IgM | 226/261 (86.5%) |
| Chikungunya | Blood | ELISA IgG | 45/271 (16.6%) |
| Chikungunya | Blood | DPP RDT IgM | 106/495 (21.4%) |
| Chikungunya | Blood | DPP RDT IgG | 55/495 (11.1%) |
| Chikungunya | Blood | DPP RDT IgM^§^ | 135/200 (67.5%) |
| Chikungunya | Blood | DPP RDT IgG^§^ | 70/200 (35%) |
| Dengue | Blood | RT-PCR | 0/294 (0%) |
| Dengue | Blood | ELISA IgM | 237/271 (87.4%) |
| Dengue | Blood | ELISA IgG | 245/261 (93.8%) |
| Dengue | Blood | DPP RDT IgM | 87/495 (17.5%) |
| Dengue | Blood | DPP RDT IgG | 353/495 (71.3%) |
| Dengue | Blood | DPP RDT IgM^§^ | 120/200 (60%) |
| Dengue | Blood | DPP RDT IgG^§^ | 133/200 (66.5%) |
| Zika | Blood | RT-PCR | 0/294 (0%) |
| Zika | Blood | ELISA IgM | 49/261 (18.7%) |
| Zika | Blood | ELISA IgG | 198/261 (75.8%) |
| Zika | Blood | DPP RDT IgM | 35/495 (7%) |
| Zika | Blood | DPP RDT IgG | 215/495 (43.4%) |
| Zika | Blood | DPP RDT IgM^§^ | 85/200 (42.5%) |
| Zika | Blood | DPP RDT IgG^§^ | 88/200 (44%) |
| HIV | Blood | Antibody detection | 7/500 (1.4%) |
| Bacteria |  |  |  |
| *Streptococcus pyogenes* | Throat | Antigen detection | 40/138 (28.9%) |
| *Streptococcus pneumoniae* | Blood | Culture | 1/498 (0.2%) |
| *Streptococcus pneumoniae* | Urine | Antigen detection | 7/135 (5.1%) |
| *Shigella flexnerii* | Stool | Culture | 1/9 (11.1%) |
| *Salmonella spp.* | Stool | Culture | 1/9 (11.1%) |
| *Salmonella spp.* | Blood | Antigen detection | 5/110 (4.54%) |
| *Escherichia coli* | Urine | Culture | 10/26 (38.4%) |
| *Escherichia coli* | Blood | Culture | 2/498 (0.4%) |
| *Klebsiella pneumoniae* | Urine | Culture | 1/26 (3.8%) |
| *Proteus mirabilis* | Urine | Culture | 1/26 (3.8%) |
| *Leptospira spp.* | Blood | Serology | 15/359 (4.1%) |
| *Rickettsia spp.* | Blood | RT-PCR | 0/358 (0%) |
| *Treponema pallidum* | Blood | Antibody detection | 30/499 (6%) |
| Parasites |  |  |  |
| *Plasmodium spp.* | Blood | Antigen detection | 0/500 (0%) |
| *Cryptococcus neoformans* | Blood | Antigen detection | 0/344 (0%) |

COR43 denotes Coronavirus 43; COR63 Coronavirus 63; COR229 Coronavirus 229; DPP RDT diagnostic test based on dual path platform technology; HADV Human Adenovirus, HBoV Human Bocavirus; HKU Coronavirus Hong Kong; HMPV A/B Human Metapneumovirus Type A/B; HPeV Human Parechovirus; HPIV1 Human Parainfluenza Type 1; HPIV2 Human Parainfluenza Type 2; HPIV3 Human Parainfluenza Type 3; HPIV4 Human Parainfluenza Type 4; HRSV A/B Human Respiratory Syncytial Virus Type A/B; NP nasopharynx; RT-PCR reverse transcription polymerase chain reaction

¶ Ten participants had more than one respiratory virus positive in a NP sample

§ Follow-up samples

Supplementary Table 3. Demographic, clinical and laboratory characteristics of study participants by the main etiologic diagnosis, Rio de Janeiro, Brazil, October 25, 2018 – July 31, 2019

| Characteristics | Chikungunya  (N=100) | Salmonellosis  (N=6) | Leptospirosis  (N= 15) | Viral URTI  (N= 40) | Pyelonephritis  (N=14) |
| --- | --- | --- | --- | --- | --- |
| Demographic variables |  |  |  |  |  |
| Female sex – no. (%) | 37 (37) | 3 (50) | 3 (20) | 24 (60) | 13 (**92.9**) |
| Age, mean (±SD) | 32.6 (16.2) | 46.8 (17.6) | 35.4 (15.3) | 18.6 (13.4) | 26.8 (**13.7**) |
| Age (years) – no. (%) |  |  |  |  |  |
| ≤ 15 | 18 (18) | 0 (0) | 2 (13.3) | 19 (47.5) | 2 (14.3) |
| 16 – 26 | 18 (18) | 1 (16.7) | 3 (20) | 10 (25) | 8 (57.1) |
| 27 - 40 | 32 (32) | 1 (16.7) | 4 (26.7) | 9 (22.5) | 2 (14.3) |
| ≥ 41 | 32 (32) | 4 (66.7) | 6 (40) | 2 (5) | 2 (14.3) |
| Clinical variables |  |  |  |  |  |
| BMI, median (IQR) | 27.6 (23.7-31.4) | 31.7 (29.2-38.5) | 28.4 (23.2-29.5) | 21.9 (18-26) | 26.1 (22-30.2) |
| Mild upper arm circumference (cm), mean (±SD) | 215.4 (59.2) | - | 210 (200-300) | 130 (90-180) | - |
| Illness duration, median (IQR) | 2 (1-3) | 2.5 92-6) | 2 (1-3) | 3 (2-4.5) | 3 (2-4) |
| Symptoms |  |  |  |  |  |
| Headache – no. (%) | 90 (90) | 5 (83.3) | 13 (86.7) | 27 (67.5) | 12 (85.7) |
| Photophobia – no. (%) | 38 (38) | 2 (33.3) | 3 (20) | 11 (27.5) | 2 (14.3) |
| Sore throat – no. (%) | 15 (15) | 0 (0) | 2 (13.3) | 27 (67.5) | 0 (0) |
| Rhinorrhea – no. (%) | 9 (9) | 1 (16.7) | 4 (26.7) | 26 (65) | 1 (7.1) |
| Cough – no. (%) | 20 (20) | 1 (16.7) | 1 (6.7) | 25 (62.5) | 2 (14.3) |
| Dysuria – no. (%) | 6 (6) | 0 (0) | 0 (0) | 3 (7.5) | 11 (78.6) |
| Diarrhea – no. (%) | 17 (17) | 2 (33.3) | 3 (20) | 5 (12.5) | 1 (7.1) |
| Joint pain – no. (%) | 88 (**88**) | 3 (50) | 10 (66.7) | 5 (12.5) | 2 (14.3) |
| Rash -no. (%) | 54 (**54**) | 1 (16.7) | 6 (40) | 3 (7.5) | 0 (0) |
| Medical past history |  |  |  |  |  |
| Recent antibiotic use – no. (%) | 3 (3) | 1 (16.7) | 3 (20) | 6 (15) | 1 (7.1) |
| Comorbidities – no. (%) | 24 (24) | 3 (50) | 5 (33.3) | 10 (25) | 2 (14.3) |
| Vital signs |  |  |  |  |  |
| Temperature (C), mean (±SD) | 37.8 (1.1) | 37.3 (0.6) | 38 (0.8) | 37.5 (1.1) | 37.6 (0.8) |
| Heart rate(beats/min), mean (±SD) | 99.1 (19.6) | 91.5 (24) | 101.8 (17.1) | 106.1 (24.4) | 112.2 (22.2) |
| Respiratory rate (rate/min), median (IQR) | 21.50 (20-24) | 18 (16-20) | 20 (18-24) | 22 (19.5-24) | 23 (21-24) |
| Mean arterial pressure (mmhg), mean (±SD) | 87.4 (25) | 96.6 (19.5) | 91.7 915.6) | 70 (34) | 88.8 (9) |
| Laboratory variables |  |  |  |  |  |
| WBC (10^9^ uL, median (IQR) | **5.7 (4.3-7.1)** | 9.4 (7.2-10.9) | 7.91 (5.5-11.7) | 8.64 (6.3-10.7) | 13.9 (9.6-17.8) |
| Lymphocytopenia (10^9^ uL) – no. (%) | 76 (76) | 3 (50) | 12 (80) | 14 (35.5) | 5 (35.7) |
| Platelets (uL), median (IQR) | 173.000 (204.000-237.000) | 230.000 (160.000-286.000) | 174.000 (154.000-220.000) | 297.500 (235.500-245.000) | 223.500 (336 -271.000) |
| Creatinine (mg/dl), median (IQR) | 0.91 (0.76-1.14) | 0.9 (0.87-1) | 1.06 (0.9-1.1) | 0.7 (0.5-0.8) | 0.81 (0.6-0.8) |
| Total bilirubin (mg/dl), median (IQR) | 0.40 (0.29-0.48) | 0.35 (0.3-0.4) | 0.4 (0.4-0.8) | 0.3 (0.2-0.5) | 0.7 (0.7-1.5) |
| CRP > 5 mg/dl – no. (%) | 59 (59) | 3 (50) | 9 (60) | 29 (72.5) | 11 (**78.6**) |

Supplementary Figure 1. Distribution of the main febrile syndromes in participants with acute febrile illness by week of illness onset, Rio de Janeiro, Brazil, October 2018-July 2019

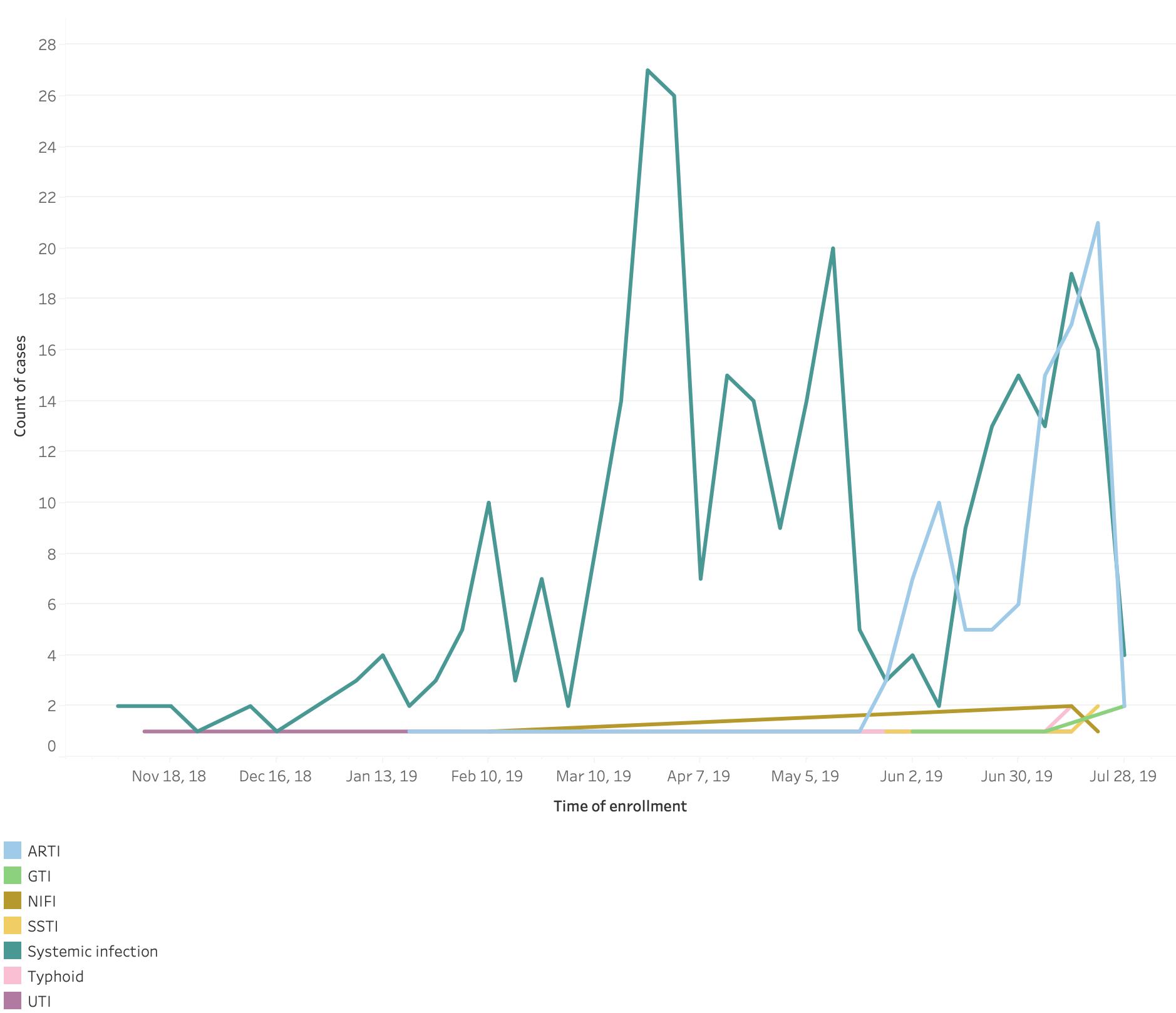

ARTI stands for acute respiratory tract infection; GTI gastrointestinal infection; NIFI non-infectious febrile illness; SSTI skin and soft tissue infection; UTI urinary tract infection

No arbovirus (n=98)

Supplementary Figure 2. Venn diagram showing the distribution of the ELISA arboviruses‘ results in the sample cohort and the proportion of co-infection between the arboviruses, Rio de Janeiro, Brazil, October 2018-July 2019

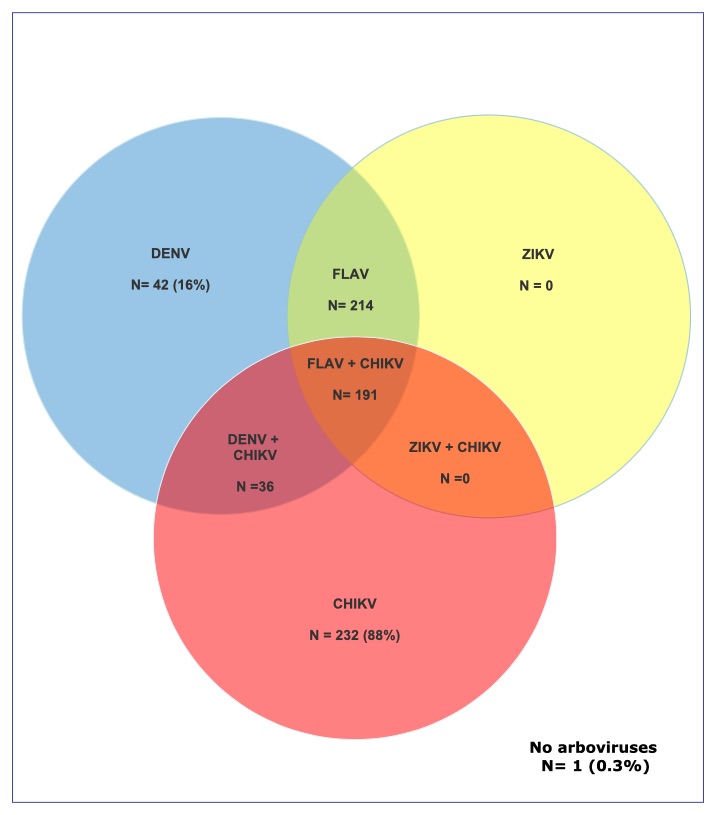

DENV means dengue virus; CHIKV chikungunya; FLAV flaviviruses; ZIKV zika virus

Supplementary Figure 3. Distribution of acute febrile illness patients according to RT-PCR evidence for chikungunya virus infection, Rio de Janeiro, October 2018-July 2019

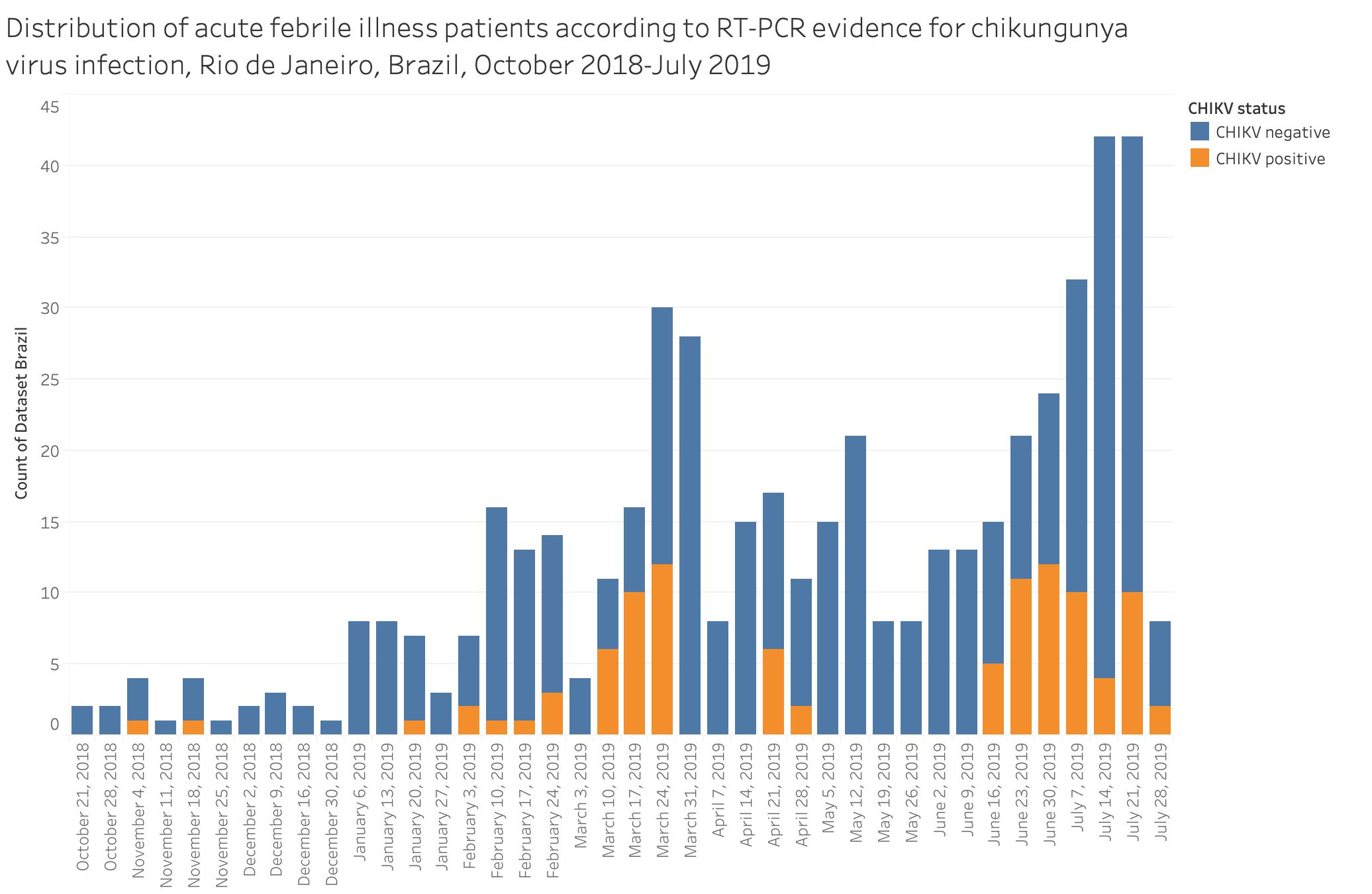

**Methods**

**Dual-Path immunochromatographic Platform rapid diagnostic test against zika, dengue, and chikungunya**

The rapid diagnostic test kits used were based on the dual-path immunochromatographic platform for IgM and IgG against ZIKV, CHIKV and DENV. The rapid diagnostics were developed by Bio-Manguinhos, Fundacao Oswaldo Cruz, Brazil, in partnership with Chembio Diagnostic System INC, USA and used whole blood, serum or plasma, digital or venous puncture samples for the simultaneous detection of IgM and IgG against the three arboviruses most abundant in Brazil. Results become available in 15 to 20 minutes. The test has an innovation that is a digital instrument for reading, interpreting and storing test results. Results were available in absolute numbers and in the form of categories based on pre-established cut-offs (reactive, non-reactive, indetermined and invalid). The manufacture test performance indicated sensitivity (IgM and IgG) close to 100% and specificity of 95% for IgM and 98% for IgG, but high level of cross reaction between ZIKV and DENV were reported.

We ran the test using participant’ serum aliquots that were stored at -80 °C, in the initial and convalescent samples.
